# Prospective multi-omic profiling identifies predictive and genetically supported molecular determinants of incident epilepsy

**DOI:** 10.64898/2026.11.18.26364966

**Authors:** Emadeldin Hassanin, Maryam Erfanian Omidvar, Priyanshu Panda, Holger Lerche, Costin Leu, Dennis Lal, Patrick May

## Abstract

Epilepsy affects approximately 50 million people worldwide, yet biomarkers capable of identifying individuals at risk before clinical onset are lacking, and the molecular mechanisms preceding disease onset remain poorly understood. Here, we integrated plasma proteomic, metabolomic, and genomic data from the UK Biobank to characterize molecular changes preceding epilepsy and evaluate their predictive and causal relevance.

We quantified 2,923 plasma proteins in ∼53,000 participants and followed incident epilepsy for up to 15 years before epilepsy diagnosis. Cox proportional hazards analyses identified 116 proteins associated with incident epilepsy, led by neurofilament light chain (NEFL; HR=2.03) and growth differentiation factor 15 (GDF15; HR=1.73). These proteins were enriched for extracellular signaling, immune regulation, and cell-death pathways and displayed distinct subtype-and age-specific signatures. A proteomic risk score comprising 32 proteins stratified epilepsy risk, with individuals in the highest quintile exhibiting a 3.5-fold greater risk than those in the lowest quintile (AUC=0.70). In parallel, 154 of 249 circulating metabolites were associated with incident epilepsy, with the strongest inverse association observed for citrate (HR=0.72) and positive associations across HDL-related metabolites (HR=1.16–1.21), implicating dysregulated energy and lipoprotein metabolism. A metabolomic risk score showed more modest discrimination (AUC=0.63). Integrating proteomic, metabolomic, polygenic, and demographic information yielded an overall AUC of 0.71 for incident epilepsy; in a separate cross-sectional analysis of individuals with early-onset epilepsy discrimination reached an AUC of 0.79.

Proteome-wide cis-pQTL Mendelian randomization identified 12 proteins associated with epilepsy after false discovery rate correction, predominantly for genetic generalized epilepsy. O-GlcNAcase (OGA), a regulator of protein O-GlcNAcylation with established roles in neuronal function and synaptic plasticity, showed the strongest protective association (OR=0.71, FDR=2.7×10⁻⁸). Serine protease 53 (PRSS53), a poorly characterized serine protease, showed strong evidence for a potential shared causal variant with epilepsy (colocalization PP.H4=0.95).

Together, these findings demonstrate that circulating molecular alterations precede epilepsy diagnosis by years and reveal complementary proteomic, metabolic, and genetic determinants of disease susceptibility, providing a framework for biologically informed risk stratification and therapeutic target prioritization.

## Introduction

Epilepsy comprises a heterogeneous group of neurological disorders characterized by a persistent predisposition to generate unprovoked seizures and affects approximately 50 million individuals worldwide, contributing significantly to the global disease burden.^1,2^ Despite major advances in neuroimaging, electrophysiology, and molecular genetics, epilepsy diagnosis remains largely symptom-driven and frequently occurs only after seizure onset. Early identification of individuals at increased risk of developing epilepsy could facilitate preventive interventions, improve disease management, and ultimately enable more personalized therapeutic strategies^3^. Consequently, identifying objective molecular biomarkers that capture biological processes preceding clinical disease onset remains a major unmet need.^4,5^

Epileptogenesis is a complex and dynamic process involving interactions between inherited genetic susceptibility, environmental exposures, neuroinflammation, metabolic dysregulation, neuronal injury, and alterations in neuronal signaling.^6–7^ Individual molecular layers provide only a partial view of the biological complexity. Recent advances in high-throughput molecular profiling now enable comprehensive characterization of complementary biological processes before disease onset. Circulating plasma proteomics reflects immune activation, neuronal injury, extracellular signalling, and tissue-specific responses, whereas plasma metabolomics captures alterations in intermediary metabolism, mitochondrial function, oxidative stress, lipid metabolism, and energy homeostasis.^8–10^ Integrating these complementary molecular layers therefore offers a unique opportunity to characterize systemic biological changes that precede epilepsy development.^11–13^

Large population-based studies have demonstrated that circulating molecular profiles can predict future disease years before clinical diagnosis. Plasma proteomics has successfully identified biomarkers associated with incident cardiovascular disease, neurodegenerative disorders, inflammatory bowel disease, and multiple chronic diseases, while recent proteogenomic studies have further elucidated the genetic architecture of the circulating proteome and its relationship to human disease.^14–16^ Likewise, metabolomic profiling has revealed predictive signatures across neurological and cardiometabolic disorders, highlighting its ability to capture dynamic biological processes that are not reflected by inherited genetic variation.^17,18^ In epilepsy, perturbations of energy metabolism are biologically plausible because the tricarboxylic acid (TCA) cycle intermediate citrate links mitochondrial function to glutamate and γ-aminobutyric acid (GABA) synthesis, while loss-of-function variants in the citrate transporter gene SLC13A5 cause developmental and epileptic encephalopathy (DEE).^19–21^ Nevertheless, the circulating metabolome has not been systematically examined in relation to incident epilepsy.

Within epilepsy, biomarker research has expanded considerably during recent years. Circulating neurofilament light chain (NEFL), glial fibrillary acidic protein (GFAP), inflammatory mediators, and several metabolic pathways have been associated with seizure burden, neuronal injury, pharmacoresistance, and disease progression.^22–26^ More recently, Zhang *et al.* demonstrated in a large prospective population-based study that plasma proteomic profiles are associated with incident epilepsy and identified molecular alterations preceding clinical disease onset, highlighting pathways related to immune regulation, inflammation, extracellular matrix remodeling, and cellular stress responses.^27^ While these findings established plasma proteomics as a promising approach for epilepsy biomarker discovery, they also raise the question: which circulating molecules merely predict future epilepsy and which are likely to contribute directly to disease pathogenesis. Furthermore, the complementary contributions of inherited genetic susceptibility, circulating proteins, and metabolic alterations to epilepsy risk remain largely unexplored.

Human genetics provides an orthogonal perspective on disease biology because germline genetic variants are fixed throughout life and are therefore largely unaffected by disease progression or reverse causation. Polygenic scores (PRSs) aggregate the effects of thousands of common variants and have emerged as a powerful tool for quantifying genetic liability across complex diseases, including generalized genetic epilepsy (GGE) and focal epilepsy (FE). However, PRSs capture inherited susceptibility rather than downstream biological processes and their predictive performance depends strongly on phenotype definition, ancestry, and the underlying genetic architecture.^28–31^ Conversely, circulating molecular profiles integrate both inherited and acquired biological processes occurring during disease development. Combining these complementary layers has already improved prediction and biological interpretation in several complex diseases, including coronary artery disease and type 2 diabetes.^32,33^

Beyond risk prediction, human genetics can also prioritize molecular mechanisms. Mendelian randomization (MR) exploits naturally occurring genetic variation to distinguish proteins that are more likely to contribute causally to disease from proteins that represent downstream responses, while Bayesian colocalization (BC) evaluates whether protein quantitative trait loci (pQTLs) and disease-associated variants share the same underlying causal signal.^34–36^ Integrating prospective molecular profiling with these genetic approaches therefore provides an opportunity to separate predictive biomarkers from genetically supported molecular determinants of epilepsy.

Here, we integrated prospective plasma proteomics, metabolomics, PRSs, and proteome-wide MR within the UK Biobank (UKB) to characterize molecular alterations preceding incident epilepsy. We first identified circulating proteins and metabolites associated with future epilepsy risk, then evaluated their predictive performance individually and in combination with inherited genetic susceptibility, and finally prioritized genetically supported molecular determinants using MR and BC (Figure 1). By integrating complementary molecular layers with human genetics, our study independently confirms and substantially extends recent prospective proteomic analyses of epilepsy by moving beyond biomarker discovery toward distinguishing predictive biomarkers from putatively causal disease mechanisms, thereby providing a comprehensive framework for biologically informed risk stratification, mechanistic discovery, and improving the prevention and management of epilepsy.

**Figure 1:**
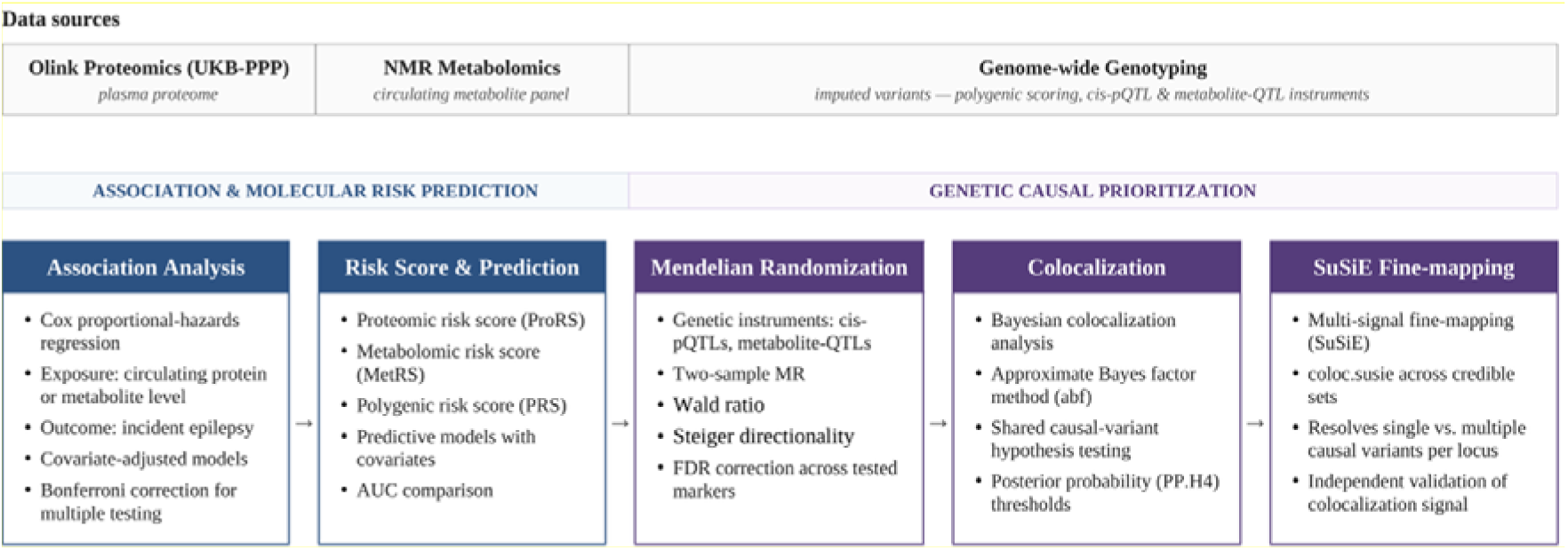
Study workflow of integrating proteomic, metabolomic, and genetic data in epilepsy. Overview of the analytical pipeline combining three data sources - Olink plasma proteomics (UKB-PPP), NMR-based circulating metabolomics, and genome-wide imputed genotyping – in epilepsy. The workflow proceeds through two linked stages. In the association and molecular risk prediction stage, Cox proportional-hazards models tested circulating protein and metabolite levels against incident epilepsy, followed by construction and comparison of proteomic (ProRS), metabolomic (MetRS), and polygenic (PRS) risk scores for predictive performance. In the genetic causal prioritization stage, cis-pQTLs and metabolite QTLs served as instruments for two-sample Mendelian randomization (Wald ratio, with Steiger filtering and FDR correction), and markers surviving correction were carried forward to Bayesian colocalization and SuSiE-based multi-signal fine-mapping to confirm a shared causal variant between molecular trait and epilepsy risk. This staged design allows observational associations to be distinguished from genetically supported causal relationships.

## Materials and methods

### Study cohort

This study utilized data from the UK Biobank (UKB), a prospective population-based cohort of ∼500,000 individuals aged 40–69 at recruitment between 2006 and 2010. Participants underwent extensive baseline phenotyping, including demographic, lifestyle, clinical, genetics, and molecular assessments, and were subsequently followed longitudinally through linked hospital admission records, death registries, and primary care data. All participants provided written informed consent. UKB received ethical approval from the North West Multi-centre Research Ethics Committee (MREC) and all analyses were performed under the relevant UKB application in accordance with the Declaration of Helsinki.^37^

### Definition of epilepsy phenotypes

Epilepsy diagnosis was defined using linked hospital records and self-reported medical history. Cases were defined by the presence of epilepsy diagnosis recorded using ICD-10 codes G40 and G41, ICD-9 code 345, or self-reported epilepsy (UKB Data-Field 20002, code 1264).^38^

Generalized genetic epilepsy (GGE) was defined using ICD-10 codes G40.3, G40.4, G40.6, and G40.7 and the corresponding ICD-9 codes 345.0-345.3. Focal epilepsy (FE) was defined using ICD-10 codes G40.0, G40.1 and G40.2 with corresponding ICD-9 345.4, 345.5, and 345.7. Participants without any epilepsy diagnosis recorded at baseline and at any follow-up were classified as controls.

### Definition of incident recorded diagnosis of epilepsy

The primary analyses focused on incident epilepsy. Incident epilepsy cases were defined as participants who received their first recorded epilepsy diagnosis after the baseline blood collection used for plasma proteomic and metabolomic profiling. Individuals with an epilepsy diagnosis preceding baseline assessment were considered prevalent cases and excluded from the primary analyses. This study design ensured that circulating molecular profiles preceded clinical disease onset, allowing evaluation of biomarkers associated with future epilepsy risk rather than established disease.

### Plasma proteomics

Plasma proteomic profiles were generated within the UKB Pharma Proteomics Project (UKB-PPP) using the Olink Explore 3072 platform based on proximity extension assays (PEA), enabling multiplexed protein quantification of 2,923 unique proteins across approximately 53,000 participants.^34^

Participants with > 50% missing protein measurements (n=3,553) and proteins >50% missing values (n=3) were excluded. Remaining missing values were imputed using the k-nearest neighbors (k=10) method with the R package *impute*.^13^ Protein abundances were analyzed as normalized protein expression (NPX) values provided by UKB-PPP.

### Plasma metabolomics

Plasma metabolomic data were generated using the Nightingale Health high-throughput nuclear magnetic resonance (NMR) metabolomics platform, which quantifies 249 circulating metabolites spanning lipoprotein subclasses, fatty acids, amino acids, ketone bodies, glycolysis-related metabolites, and other intermediates.^18,39^

Participants with >50% missing in metabolite measurements were excluded. The remaining missing values were imputed using the k-nearest neighbors imputation (k=10). Metabolite concentrations were subsequently log-transformed to reduce skewness and standardized (mean = 0, SD = 1) prior to downstream association analyses and risk score modeling.

### Prospective association analysis

Associations between baseline plasma protein abundance and incident epilepsy were evaluated using Cox proportional hazards regression models, with protein normalized protein expression (NPX) values included as continuous predictors. Hazard ratios (HRs) and corresponding 95% confidence intervals (CIs) were estimated for each protein. All models were adjusted for age at assessment, sex, body mass index (BMI), smoking status, alcohol consumption, history of stroke, and the first four genetic principal components (PCs) to account for population stratification. To account for multiple testing across all proteins, statistical significance was determined using Bonferroni correction, with a Bonferroni-adjusted P-value < 0.05 considered significant.

To evaluate whether identified biomarkers were also associated with established disease, we performed secondary logistic regression analyses of prevalent epilepsy, GGE and FE using the same covariates. Additional subgroup analyses stratified cases by age at diagnosis into childhood-onset (<10 years), juvenile-onset (10-18 years), and adult-onset (>18 years) groups. Associations are reported as odds ratios (ORs) with 95% CIs. Sex-stratified Cox regression analyses were additionally performed to identify sex-specific prospective associations.

Associations between baseline metabolite concentrations and incident reported epilepsy were assessed in the full eligible UK Biobank dataset with available metabolomics measurements, including 3,418 incident reported epilepsy cases, using the same Cox regression framework and covariate adjustment as the proteomic analyses. Bonferroni correction was applied across all 249 metabolites.

### Construction of molecular risk scores

A proteomic risk score (ProRS) for incident epilepsy was constructed using Least Absolute Shrinkage and Selection Operator (LASSO) regression implemented via the *glmnet* R package. LASSO simultaneously performs variable selection and coefficient regularization, making it particularly well suited for high-dimensional molecular datasets by shrinking non-informative coefficients toward zero while retaining most predictive features. Prior to model fitting, NPX values were standardized (mean = 0, SD = 1).

The dataset was randomly divided into equally sized training and testing subsets. Cox LASSO regression was used, adjusting for age, sex, and the first four genetic principal components (PCs). Missing-value imputation and standardization were performed within the training data and cross-validation folds to prevent information leakage, and the optimal λ was selected using five-fold cross-validation. The final model was refitted on the training set and applied unchanged to the independent testing set to calculate individual-level ProRS.

A metabolomic risk score (MetRS) for incident epilepsy was generated using the same analytical framework. Standardized metabolite concentrations were used as input, the dataset was split into equally sized training and testing subsets, and the covariate-adjusted LASSO regression with five-fold cross-validation was performed to identify the optimal λ before calculating MetRS values for the independent test cohort.

### Construction of polygenic risk scores

Polygenic risk scores (PRSs) for epilepsy were computed for UKB participants using the summary statistics from the latest genome-wide association study (GWAS Catalog: GCST90271611) from the International League Against Epilepsy (ILAE) Consortium on Complex Epilepsies (GWAS).^30^ We used the ILAE summary statistics that did not include UKB participants in the meta-analysis, thereby ensuring independence between the discovery GWAS and the UKB target dataset and minimizing potential overfitting and sample overlap.

PRS were generated using PRS-CS,^40^ a Bayesian regression framework that infers posterior SNP effect sizes using continuous shrinking priors while accounting for local linkage disequilibrium (LD) patterns based on an external reference panel (1000 Genomes). Compared with conventional clumping-and-thresholding approaches, PRS-CS jointly models genome-wide marker effects and typically improves prediction for highly polygenic traits. The resulting PRSs were standardized (mean = 0, SD = 1) prior to downstream analyses and subsequently evaluated both individually and in combination with the ProRS and MetRSs.

### Functional and pathway enrichment analysis

Functional enrichment analysis was performed to characterize biological pathways and processes represented by proteins significantly associated with incident epilepsy in the Cox proportional hazards analyses (Bonferroni-adjusted P<0.05). Gene set enrichment was performed using g:Profiler,^41^ incorporating annotations from Gene Ontology (GO), Kyoto Encyclopedia of Genes and Genomes (KEGG), and Reactome pathway databases. Enrichment significance was assessed using g:Profiler’s default multiple-testing correction, and significantly enriched terms were ranked according to adjusted *P*-values. Significantly enriched pathways were ranked according to adjusted *P*-values, and the four most significant terms from each annotation database were visualized.

### Genetic prioritization of putatively causal proteins

To distinguish proteins that may contribute causally to epilepsy development from proteins representing downstream disease-associated biological processes, we performed two-sample Mendelian randomization (MR) using independent cis-protein quantitative trait loci (cis-pQTL) as instrumental variables.

Exposure data were obtained from the UKB-PPP, which quantified 2,923 plasma proteins in 54,219 individuals and identified 14,287 significant cis-pQTLs after multiple-testing correction.^34^ Genetic instruments were required to satisfy the following criteria: (i) genome-wide significance (*P* < 5 × 10⁻⁸), (ii) localization outside the major histocompatibility complex (MHC; chromosome 6: 26–34 Mb), (iii) independence following LD clumping (*r²* < 0.001), (iv) localization within ±1 Mb of the corresponding protein-coding gene, and (v) an F-statistic >30.

Epilepsy GWAS summary statistics from the ILAE Consortium^30^ served as the outcome dataset. Because all proteins were instrumented by a single independent cis-pQTL, causal effects were estimated using the Wald ratio. Instrument validity was evaluated using the F-statistic and the Steiger directionality test,^35^ and false discovery rate (FDR) correction was applied across all tested proteins within each epilepsy outcome.

For proteins remaining significant after FDR correction, Bayesian colocalization was performed using the approximate Bayes factor method (*coloc.abf*) implemented in the coloc R package.^36^ Analyses were conducted within ±500 kb of the lead cis-pQTL using default prior probabilities. Posterior probabilities for hypothesis 4 (PP.H4) were interpreted as evidence for a shared causal variant, with PP.H4 ≥ 0.80 indicating strong evidence for colocalization, PP.H4 = 0.50–0.79 suggestive evidence, and PP.H4 < 0.50 indicating little or no evidence of a shared signal.

To account for loci containing multiple independent association signals, we additionally performed multi-signal colocalization using the SuSiE fine-mapping framework.^42^ Exposure and outcome datasets were independently fine-mapped prior to *coloc.susie* analysis, and the highest PP.H4 across all credible-set pairs was reported as a sensitivity analysis that relaxes the single-causal-variant assumption.

### Metabolite Mendelian randomization

To evaluate whether circulating metabolic traits contribute causally to epilepsy risk, we performed a parallel two-sample MR analysis using metabolite quantitative trait loci (mQTLs) as instrumental variables. Exposure data were obtained from a UKB NMR metabolomics fine-mapping study comprising 249 Nightingale metabolic traits, in which fine-mapped credible sets generated using SuSiE and supported by FINEMAP were represented by their highest posterior inclusion probability (PIP) variants.^43^

Genetic instruments were required to reach genome-wide significance (*P* < 5 × 10⁻^8^) and an F-statistic >10. Within each metabolite, correlated variants (*r²* ≥ 0.001) were pruned using a European 1000 Genomes reference panel, retaining at least five independent instruments per metabolite. GWAS summary statistics from the ILAE Consortium for all epilepsy, GGE), and FE served as outcome datasets.^30^

Inverse-variance weighted (IVW) Mendelian randomization was used as the primary analysis, with MR-Egger, weighted median, and weighted mode serving as sensitivity analyses. Heterogeneity and horizontal pleiotropy were assessed using Cochran’s Q statistic, the MR-Egger intercept, and MR-PRESSO. Instrument orientation was confirmed using Steiger filtering. False discovery rate correction was applied across all metabolites within each outcome. Because no metabolite remained significant after multiple-testing correction, colocalization analyses were not performed.

### Integration of genetic and molecular risk scores

To evaluate the complementary predictive value of genetic and molecular biomarkers for incident epilepsy, we systematically compared ten prediction models incorporating combinations of polygenic (PRS), proteomics (ProRS), and metabolomic (MetRS) risk scores.

Ten logistic regression models were evaluated, including a baseline model containing age, sex, and the first four genetic principal components; models including each individual risk score; baseline-adjusted models; pairwise combinations of risk scores; and a fully integrated model combining PRS, ProRS, MetRS, and demographic covariates. A complete description of all prediction models is provided in Supplementary Table S1.

For analyses of epilepsy onset at ≤18 years, because UK Biobank blood samples were collected at baseline when participants were aged 40–69 years, the molecular measurements were obtained years after disease onset and were therefore treated as cross-sectional sensitivity analyses, separate from the prospective incident-epilepsy analyses.

Model discrimination was assessed using the area under the receiver operating characteristic curve (AUC) with corresponding 95% CIs, enabling direct comparison of the predictive performance of individual molecular layers and their combinations relative to the baseline model.

To evaluate clinical risk stratification, participants were classified into high-risk (>80th percentile) and low-risk (≤20th percentile) groups according to their PRS and ProRS. Cox proportional hazards regression was then used to estimate HRs and 95% CIs for incident epilepsy during follow-up.

## Results

### Cohort characteristics

A total of 49,461 UKB participants with matched plasma proteomic, plasma metabolomic, and genomic data were included in the analyses. During the prospective follow-up, 400 individuals developed incident epilepsy, while 609 participants had prevalent epilepsy at baseline and were excluded from the primary prospective analyses. Incident cases were older at baseline than controls (mean = 59.0 vs. 56.8 years) and included a slightly higher proportion of males (49.0% vs. 45.9%) (Table 1). Among incident epilepsy cases 55 (13.8%) were classified as generalized genetic epilepsy (GGE) and 31 (7.8%) as focal epilepsy (FE), while the remaining cases represented other or unspecific epilepsy subtypes.

**Table 1:** Characteristics of the participants categorized by incident epilepsy status.

|  | **Incident Cases** | **Prevalent Cases** | **Controls** |
|---|---|---|---|
| **Participants, N** | 400 | 609 | 48,452 |
| **Male, N (%)** | 196 (49 %) | 316 (51.9 %) | 22249 (45.92 %) |
| **Female, N (%)** | 204 (51 %) | 293 (48.1 %) | 26203 (54.08 %) |
| **Age, mean (SD)** | 58.98 (7.76) | 56.57 (8.4) | 56.8 (8.21) |
| **Genetic generalized epilepsy, N (%)** | 55 (13.75 %) | 80 (13.13 %) | _ |
| **Focal epilepsy, N (%)** | 31 (7.75 %) | 54 (8.86 %) | _ |

### Plasma proteome captures prospective epilepsy risk

Consistent with recent large-scale proteomic studies of epilepsy^27^, prospective plasma proteomic profiling identified 116 proteins significantly associated with incident epilepsy after Bonferroni correction, demonstrating that widespread systemic molecular alterations are detectable years before clinical diagnosis (Figure 2A; Supplementary Table S2). The majority of significant proteins showed increased abundance in individuals who subsequently developed epilepsy, consistent with early activation of inflammatory, immune, and neuronal injury pathways.

**Figure 2:**
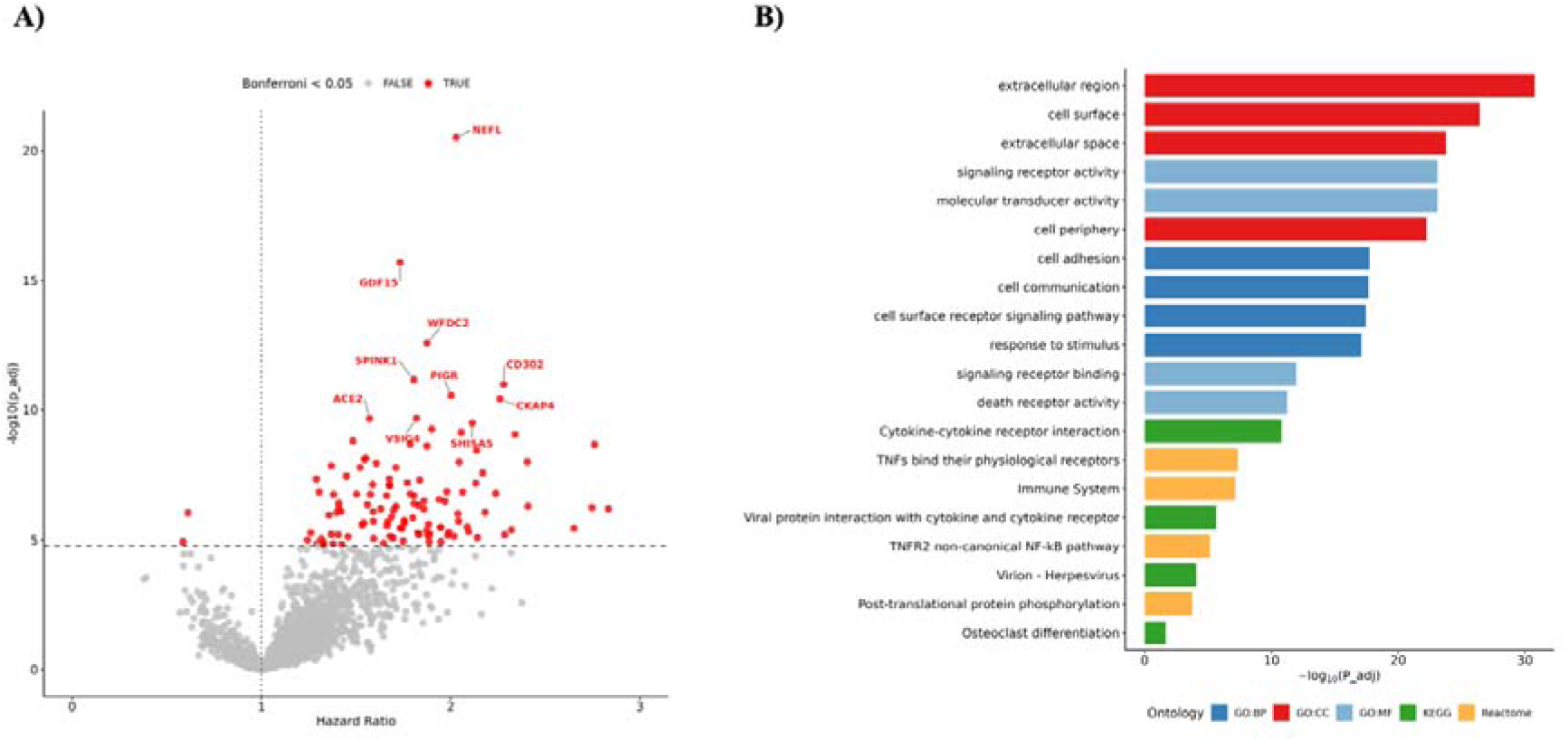
Plasma proteomic associations and pathway enrichment for incident epilepsy. A) Volcano plot summarizes the results of the Cox proportional hazards regression models. B) Functional enrichment analysis performed using gProfiler, showing the top 4 enriched terms for various ontologies (GO-BP: Gene Ontology Biological Process, GO-CC: Gene Ontology Cellular Component, GO-MF: Gene Ontology Molecular Function, KEGG, and Reactome).

The strongest association was observed for neurofilament light chain (NEFL) (HR = 2.03, *P* = 3.17 × 10⁻^21^), a well-established marker of neuroaxonal injury. Other highly significant proteins included GDF15, WFDC2, SPINK1, CKAP4, CD302, PIGR, VSIG4, ACE2, and SHISA5, representing pathways involved in cellular stress responses, innate immunity, epithelial defense, vascular regulation, and apoptosis.

Sex-stratified analyses revealed pronounced differences in the prospective plasma proteome (Supplementary Figure S1). Females exhibited substantially more significant associations than males (59 versus 13 Bonferroni-significant proteins), with 56 female-specific and 10 male-specific associations, indicating marked sex-dependent molecular signatures preceding epilepsy onset. Among females, the strongest associations were observed for WFDC2, EPHA2, and CKAP4, whereas TIMP4, BST2, and GGT1 showed the strongest associations in males.

We next investigated whether prospective biomarkers were also associated with established epilepsy. Several proteins demonstrated substantially larger effect sizes in prevalent disease than in incident epilepsy (Supplementary Figure S2), consistent with progressive amplification of inflammatory and lysosomal pathways following disease onset. Subtype analyses further identified distinct molecular signatures for generalized genetic epilepsy (GGE) and focal epilepsy (FE) (Supplementary Figure S3). GGE was characterized by numerous subtype-specific associations, including PIGR, HYAL1, ANPEP, CEACAM1, and LAMP2, whereas FE showed only four subtype-specific proteins (IL1R1, CSF1, RAB6A, and SELENOP). Likewise, age-at-onset analyses identified distinct proteomic signatures across childhood-, juvenile-, and adult-onset epilepsy (Supplementary Figure S4), suggesting that molecular pathways preceding epilepsy differ according to disease subtype and age at onset.

Together, these findings demonstrate that circulating plasma proteins capture a broad spectrum of biological alterations preceding epilepsy diagnosis with evidence for pronounced sex-, subtype-, and age-specific molecular signatures.

### Biological pathways underlying epilepsy risk

Functional enrichment analysis demonstrated that proteins associated with incident epilepsy predominantly participate in extracellular signaling, immune regulation, and receptor-mediated communication (Figure 2B). The strongest enrichments were observed for extracellular region and extracellular space components, together with signaling receptor activity and molecular transducer activity, indicating that early epilepsy-associated molecular alterations are largely mediated through secreted proteins and cell-surface signaling.

These findings are consistent with accumulating evidence implicating neuroinflammation, immune activation, and extracellular signaling as central processes during epileptogenesis.^44,45^ Rather than reflecting isolated molecular changes, the prospective proteomic signature therefore suggests coordinated activation of biological pathways involved in neuronal injury, immune regulation, and intercellular communication years before clinical disease onset.

### Mendelian randomization identifies proteins associated with epilepsy risk

To distinguish proteins that may contribute causally to epilepsy development from proteins representing downstream disease-associated biomarkers, we performed proteome-wide two-sample MR using independent cis-pQTLs as genetic instruments.

Among the 2,923 proteins evaluated, 13 protein-epilepsy associations, involving 12 unique proteins, reached significance after false discovery rate (FDR) correction, indicating that only a small subset of circulating proteins showed genetic support for a potential causal role (Figure 3, Supplemental Table S3).

**Figure 3:**
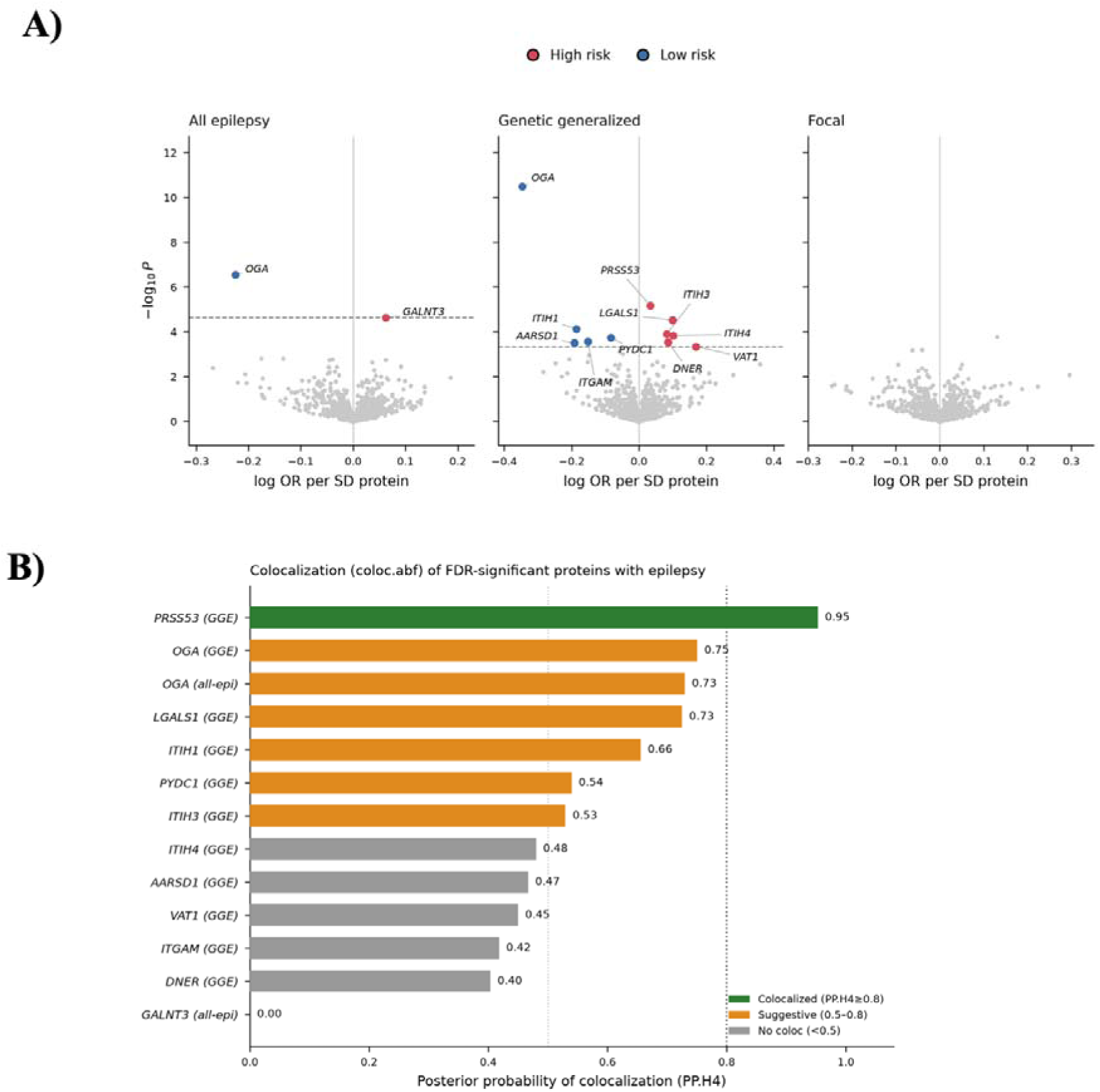
Protein-epilepsy Mendelian randomization and colocalization results. A) Volcano plot summarizes the results of proteome-wide Mendelian randomization of protein levels against all epilepsy, genetic generalized epilepsy, and focal epilepsy; labeled proteins are highlighted findings. B) Posterior probabilities of colocalization for significant proteins, with bars colored by evidence level and the 0.8 threshold marked by the dotted line.

All significant associations passed the Steiger directionality test, supporting the inferred direction of causality from genetically predicted protein abundance to epilepsy risk. The strongest association was observed for OGA, where higher genetically predicted plasma protein levels were associated with a substantially lower risk of GGE (OR = 0.71, 95% CI = 0.64–0.78, FDR = 2.7 × 10⁻⁸). OGA also showed a protective association with the ‘all epilepsy’ group (OR = 0.80, 95% CI = 0.73–0.87, FDR = 2.2 × 10⁻⁴).

Increased genetically predicted levels of PRSS53, LGALS1, ITIH3, ITIH4, DNER, VAT1, and GALNT3 were associated with increased epilepsy risk, whereas ITIH1, PYDC1, ITGAM, and AARSD1 showed protective associations (Figure 3A; Supplementary table S3). Overall, genetically supported associations were predominantly observed for GGE, consistent with its stronger genetic architecture and higher heritability.

To evaluate whether the pQTL and epilepsy GWAS signals were driven by a shared causal variant, we performed Bayesian colocalization (BC) analyses (Figure 3B). We observed a clear hierarchy of evidence across the genetically prioritized proteins. PRSS53 was the only association showing strong evidence for colocalization, with high posterior probability of a shared causal variant using both the conventional approximate Bayes factor approach (PP.H4 = 0.95) and multi-signal SuSiE fine-mapping (PP.H4 = 0.91). OGA and five additional associations showed suggestive evidence for colocalization (PP.H4 = 0.53–0.75), including OGA, LGALS1, ITIH1, ITIH3, and PYDC1 for GGE, and OGA for ‘all-epilepsy’. In contrast, six MR associations (ITIH4, ITGAM, DNER, AARSD1, VAT1, and GALNT3) showed no evidence of a shared causal variant (PP.H4 < 0.50), indicating that the observed protein–epilepsy associations may reflect distinct variants in linkage disequilibrium rather than a common causal signal.

Together these findings demonstrate that only a subset of proteins associated with prospective epilepsy risk also exhibit genetic evidence supporting a potential causal role in disease pathogenesis. The combination of MR, directionality testing, and BC therefore distinguishes predictive biomarkers from genetically supported molecular determinants, providing a framework for potentially prioritizing candidate therapeutic targets.

### Plasma metabolome captures prospective epilepsy risk

Consistent with the proteomic findings, prospective metabolomic profiling revealed widespread systemic alterations preceding epilepsy diagnosis. Among the 249 NMR-quantified metabolites, 154 were significantly associated with incident epilepsy after Bonferroni correction (Figure 4; Supplementary Table S4), suggesting that metabolic dysregulation is detectable prior to disease onset. The significant associations predominantly involved pathways related to energy metabolism, lipoprotein metabolism, ketone body metabolism, and amino acid homeostasis, suggesting broad perturbations of systemic metabolism before epilepsy develops.

**Figure 4:**
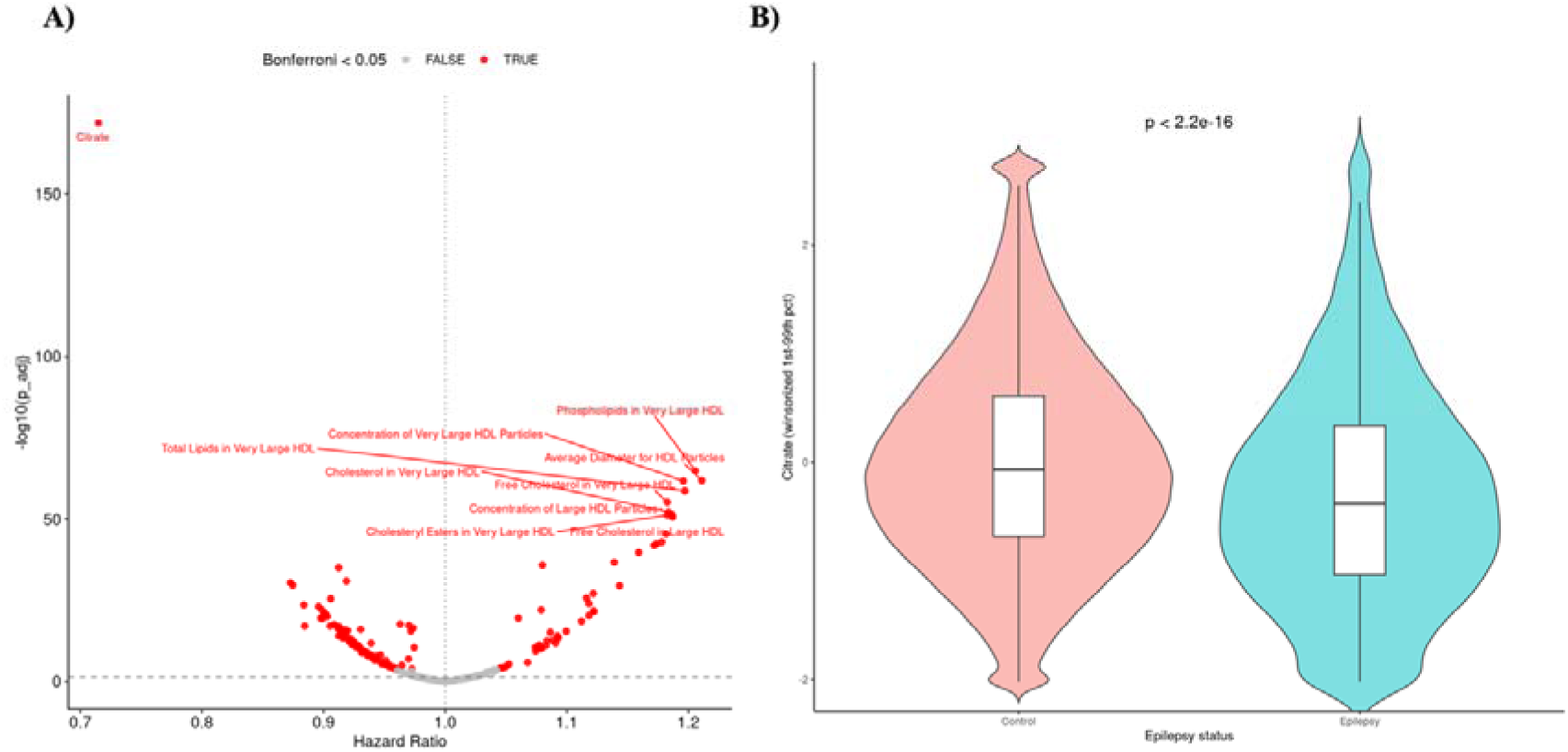
Plasma metabolome associations with incident epilepsy. A) Volcano plot summarizes the results of the Cox proportional hazards regression models. B) Distribution of serum citrate concentration in epilepsy cases versus controls.

The strongest signal was observed for citrate (HR=0.72; *P*=1.41×10⁻^172^), a central intermediate of the tricarboxylic acid (TCA) cycle, with lower circulating citrate levels associated with increased epilepsy risk. Several HDL-related measures, including phospholipids in very large HDL (HR=1.21; *P*=1.43×10⁻^65^) and mean HDL particle diameter (HR=1.21; *P*=1.30×10⁻^62^), were positively associated with incident epilepsy, whereas multiple LDL-and VLDL-associated cholesterol measures showed inverse associations (HR=0.91-0.94). Among the small-molecule metabolites, the ketone body acetone (HR=1.08; *P*=1.50×10⁻^36^) was positively associated with epilepsy risk, while the amino acid histidine showed a protective association (HR=0.87; *P*=3.48×10⁻^31^).

Notably, the observed metabolomic signatures converge on pathways involved in mitochondrial energy metabolism, lipid transport, and intermediary metabolism, complementing the inflammatory and neuronal injury signatures identified in the plasma proteome. In particular, the strong association for citrate suggests that alterations in the TCA-cycle metabolism may represent one of the earliest systemic metabolic changes preceding epilepsy diagnosis.

Two-sample MR analysis found no metabolite surviving FDR correction in any of the three epilepsy outcomes (Supplementary Figure S5).

### Comparative performance of genetic and molecular risk scores

To evaluate the predictive performance of the different molecular layers, we compared models incorporating the polygenic risk score (PRS), proteomic risk score (ProRS), metabolomic risk score (MetRS), and their combinations (Figure 5; Supplementary Table S1).

**Figure 5:**
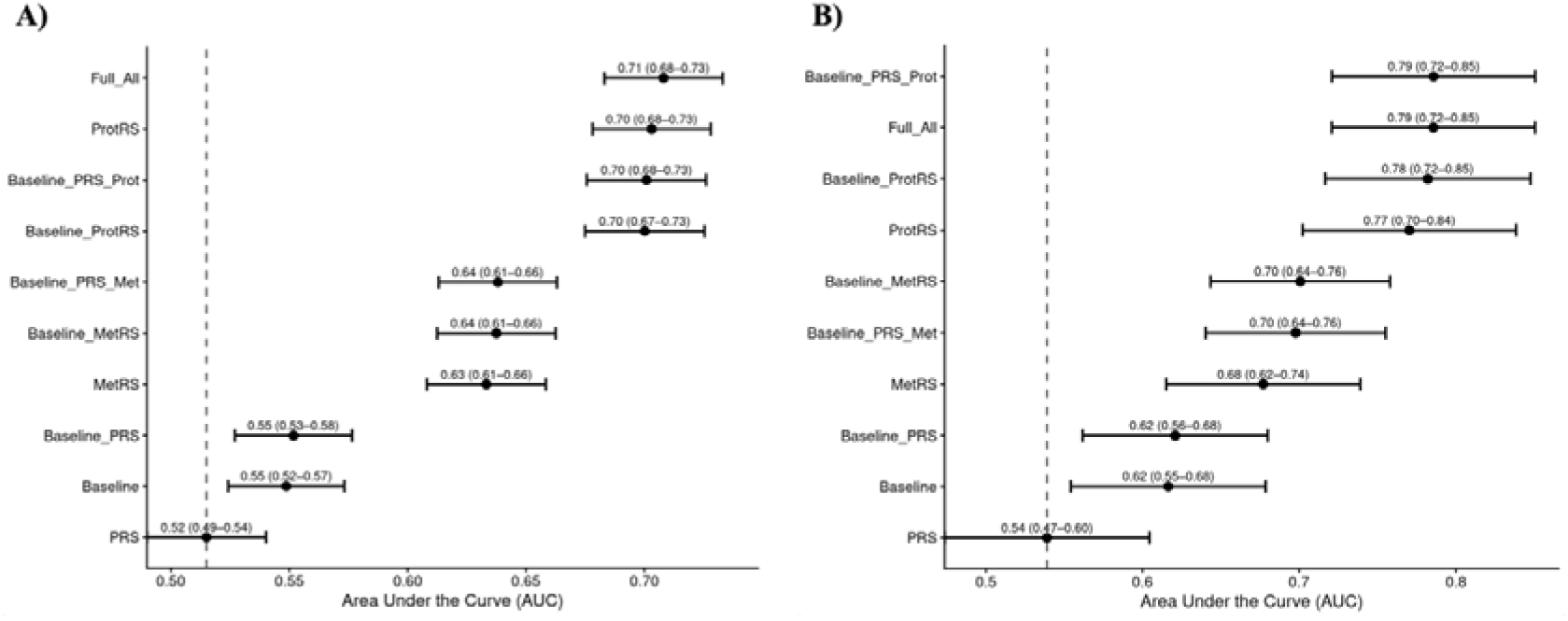
Comparison of model discrimination for incident epilepsy across polygenic, proteomic, and metabolomic risk scores. Forest plots show the area under the receiver operating characteristic curve (AUC) with 95% confidence intervals for logistic-regression models combining the polygenic (PRS), proteomic (ProRS), and metabolomic (MetRS) risk scores with demographic covariates (age, sex, and the first four genetic principal components), with models ordered by performance. Models include each score alone, each score added to the baseline covariates, pairwise combinations, and the full model integrating all three scores. (A) All epilepsy cases and controls. (B) Individuals with epilepsy onset at ≤18 years of age and controls.

Among the individual molecular predictors (Figure 5A), the ProRS achieved the highest discriminatory performance for epilepsy (AUC = 0.70, 95% CI 0.68–0.73), substantially outperforming both the MetRS (AUC = 0.63, 95% CI 0.61–0.66) and the PRS (AUC = 0.52, 95% CI 0.49–0.54). Incorporating demographic covariates resulted in only modest improvements in predictive performance, while the fully integrated model combining PRS, ProRS, MetRS, age, sex, and genetic principal components achieved the highest overall performance (AUC = 0.71, 95% CI 0.68–0.73). Prediction performance was substantially higher among individuals with epilepsy onset at ≤18 years (Figure 5B).

To further evaluate clinical risk stratification, participants were grouped according to their molecular risk scores. Individuals in the high ProRS category had a 3.5-fold increased risk of epilepsy compared with those in the lowest category (Supplementary Figure S6), whereas the PRS alone showed more limited discrimination between risk groups. These findings suggest that the ProRS may provide additional predictive value beyond common genetic risk for future epilepsy in this population.

Overall, the comparison of prediction models indicates that proteomic biomarkers provide the strongest prospective predictive signal in this population. Although metabolomic and polygenic scores captured complementary dimensions of risk, their addition resulted in only modest improvent over the ProRS alone suggesting that prospective plasma proteomics captures much of the predictive information available before clinical epilepsy onset.

### Biological architecture of the proteomic risk score

Given the superior predictive performance of the ProRS, we next examined the proteins retained by the LASSO model to characterize the biological processes underlying its discriminative ability (Supplementary Table S5). Rather than relying on a single biomarker, the ProRS integrated 32 proteins representing multiple complementary aspects of epilepsy biology, including neuronal injury, glial activation, immune regulation, lysosomal function, cellular stress responses, and vascular biology.

NEFL contributed the largest positive coefficient in the model (weight = 0.443), consistent with its role as sensitive marker of neuroaxonal injury and its strong prospective association with incident epilepsy. Other highly weighted proteins included RNF149 (0.234), CPVL (0.140), CTSL (0.135), and VSIG4 (0.115), implicating immune modulation, lysosomal activity, and neuroinflammatory pathways as major contributors to epilepsy risk prediction. Additional positively weighted proteins, including GDF15, GFAP, SPINK1, and ACE2, further highlighted the contribution of cellular stress responses, astroglial activation, and epithelial defense, and vascular regulation to the prospective molecular signature.

Conversely, RAB6A (−0.211), TNR (−0.156), and NRXN3 (−0.112) were assigned negative coefficients, indicating that higher circulating levels of these proteins contributed negatively to the prediction model. These proteins are involved in vesicle trafficking, extracellular matrix organization, and synaptic function, suggesting that preserved neuronal homeostasis may characterize individuals at lower risk of developing epilepsy.

## Discussion

Using prospective multi-omic profiling integrated with human genetics, we demonstrate that circulating plasma proteins and metabolites capture biological processes preceding epilepsy diagnosis and provide information complementary to inherited genetic susceptibility.

Our findings independently confirm and substantially extend the recent prospective plasma proteomic analysis by Zhang et al.,^27^ which similarly identified NEFL and GDF15 as the strongest circulating predictors of incident epilepsy. Our study extends this work by integrating proteomics with metabolomics, polygenic score (PRS) prediction, MR, and Bayesian colocalization (BC) to distinguish predictive biomarkers from genetically supported molecular determinants and to evaluate their complementary value for prospective epilepsy risk stratification.

Consistent with both studies, NEFL and GDF15 emerged as biologically compelling biomarkers among the 116 proteins associated with incident epilepsy. NEFL is a well-established indicator of neuroaxonal injury^46^, and has consistently been linked to seizure burden^22,23^, drug-resistant epilepsy^24,25^, cognitive impairment, supporting the concept that neuronal damage is already detectable during the preclinical phase of epilepsy.^26^ GDF15, a stress-response cytokine induced by mitochondrial dysfunction and cellular stress, has emerged as a promising biomarker across neurological disorders^47,48^. Recent studies reporting elevated GDF15 levels in status epilepticus associated with fever^49^ and in epilepsy patients with primary mitochondrial disorders^50^ further support a role for systemic mitochondrial and cellular stress responses during epileptogenesis. Beyond these individual markers, the predominance of proteins involved in immune regulation, lysosomal biology, vascular function, and glial activation suggests that epilepsy development is accompanied by coordinated systemic biological responses well before clinical onset. The pronounced sex-specific signatures observed in our study indicate that biological mechanisms preceding epilepsy may differ between males and females.

The proteomics risk score (ProRS), derived from only 32 proteins, represented the strongest predictor of incident epilepsy, stratifying individuals in the highest quintile into a 3.5-fold higher risk group than those in the lowest quintile. The ProRS outperformed both the metabolomic risk score (MetRS) and the current epilepsy PRS, while integration of additional molecular layers provided only modest improvements in discrimination. Discriminative performance was notably stronger when analyses were restricted to individuals with early-onset (<18 years) epilepsy (AUC=0.79). As molecular profiling in UKB occcured later in adulthood, this analysis should not be interpreted as prospective prediction of childhood-onset epilepsy. Rather, it suggests that the combined molecular and genetic profile more strongly distinguishes individuals with a history of early-onset epilepsy, a group expected to contain a greater proportion of genetic epilepsies. Importantly, proteomic and genetic risk contributed independently and additively, with individuals carrying both high ProRS and high PRS exhibiting the highest cumulative incidence of epilepsy. These findings are consistent with studies from other complex diseases such as stroke,^15^ Alzheimer’s,^51^ and cardiovascular disease^52^, where circulating protein signatures outperform polygenic risk scores for short-term disease prediction while capturing ongoing pathophysiological processes.

The substantially higher discriminatin achieved by the ProRS compared with the PRS should not be interpreted as indicating a limited contribution of inherited susceptibility to epilepsy. The two scores capture fundamentally different dimensions of risk; the PRS represents lifelong inhertited susceptibility, whereas circulating proteins reflect dynamic biological states at the time of sampling and may capture age-related, acquired, and prodomal processes preceding diagnosis. The superior predictive performance of the ProRS should therefore not be interpreted as evidence that all constituent proteins are causal mediators of epilepsy. Rather, circulating signatures likely integrate multiple downstream biological processes during the prodromal phase of disease. Our MR analyses support this interpretation by identifying a subset of proteins with genetic evidence consistent with potential causal involvement. Together, these complementary analyses illustrate how prospective molecular profiling and human genetics can distinguish predictive biomarkers from putative therapeutic targets.

To identify proteins more likely to contribute to disease pathogenesis, we performed proteome-wide cis-pQTL MR combined with BC and fine-mapping analyses. Twelve proteins showed genetic support for causal involvement in epilepsy, predominantly in genetic generalized epilepsy (GGE), consistent with the higher heritability and stronger contribution of common genetic variation in this subtype.^53^ Among these, PRSS53 emerged as the highest-confidence candidate, supported by convergent evidence from MR, colocalization, and SuSIE fine-mapping. Although PRSS53 has not previously been implicated in epilepsy, it belongs to a family of serine proteases with established roles in epithelial sodium channel regulation^54^ and has been associated with several neurological disorders^55,56^, making it an attractive candidate for future functional studies. Conversely, OGA represented the strongest genetically supported protective association across epilepsy outcomes. O-GlcNAcylation has increasingly been implicated in neuronal function and synaptic plasticity^57^, suggesting that altered O-GlcNAc signaling may represent another promising avenue for future mechanistic studies. More broadly, the limited overlap between predictive proteins and genetically supported proteins highlights the importance of integrating longitudinal molecular profiling with human genetics. While many circulating proteins predict future disease, only a subset shows genetic evidence consistent with potential involvement in epilepsy pathogenesis.

Subtype-specific analyses further highlighted the biological heterogeneity of epilepsy. GGE showed substantially broader proteomic dysregulation than focal epilepsy (FE), with prominent signals involving immune regulation, blood-brain barrier integrity (PIGR^58^ and CEACAM1^59^), neuroinflammation (ANPEP ^60^), and lysosomal function (LAMP2^61^). In contrast, FE was characterized by comparatively fewer associations involving IL1R1, CSF1,^62,63^ RAB6A, and SELENOP^64^, reinforcing the importance of neuroinflammatory and oxidative stress pathways in focal epileptogenesis while suggesting distinct systemic molecular responses across epilepsy subtypes. Age-at-onset analyses further identified unique molecular signatures in childhood-and juvenile-onset epilepsy. In childhood-onset cases, ATRN and LEP were uniquely significant; whereas in juvenile-onset cases, unique associations included A1BG, CFB, COL9A1, DPP4, LAG3, and TGFBI. These findings align with previous evidence linking adrenergic signaling^65^, complement activation^66^, DPP4-mediated microglial changes^67^, and TGFB-driven^68^ blood-brain barrier dysfunction to epilepsy pathophysiology. Together, these findings support the concept that molecular processes preceding epilepsy differ according to both disease subtype and age at onset, reinforcing the biological heterogeneity of epileptogenesis.

Extending the proteomic analyses, we identified 154 circulating metabolites associated with incident epilepsy, indicating that systemic metabolic remodeling also precedes clinical disease onset. The strongest association was observed for citrate, a central metabolite linking mitochondrial energy metabolism with glutamate and GABA biosynthesis^69^. Particularly noteworthy is the established role of loss-of-function variants in the neuronal citrate transporter gene *SLC13A5*, which cause an early-infantile developmental and epileptic encephalopathy^20,21^, providing a direct genetic link between citrate metabolism and seizures. Although numerous lipid and lipoprotein measures were also associated with epilepsy risk^70^, metabolomic prediction performance remained modest (MetRS AUC=0.63) and contributed little beyond the ProRS, suggesting that the circulating proteome captures a larger proportion of biologically relevant variation in this population than the metabolome.

Several limitations warrant consideration. First, although UK Biobank (UKB) offers one of the largest prospective multi-omic data sets at population scale, the relatively small number of incident epilepsy cases limited statistical power for subtype-specific analyses. Second, in UKB participants are predominantly of European ancestry and are affected by healthy volunteer bias, potentially limiting generalizability of our findings to other populations^71^. Third, epilepsy ascertainment relied primarily on hospital records using ICD coding and self-reported diagnoses, resulting in a heterogeneous mixture of epilepsy phenotypes, including GGE and FE together with structural, vascular, inflammatory, metabolic, and other acquired epilepsies. Consequently, the incident epilepsy phenotype represented in UKB differs substantially from the clinically characterized cohorts used to derive current epilepsy GWAS and PRSs. This difference in phenotype composition is likely to explain the relatively modest performance of the epilepsy PRS observed in our study. Fourth, the UKB predominantly includes middle-aged and older adults. Current epilepsy PRSs are primarily based on large case-control GWAS enriched for individuals with epilepsy onset during childhood, adolescence, or early adulthood, where inherited genetic susceptibility plays a particularly prominent role. In contrast, incident epilepsy in UKB occurs at considerably older ages and includes a substantial proportion of acquired epilepsies, for which currently available epilepsy PRSs are not expected to capture fully the underlying disease risk. This phenotype and-at-onset mismatch is likely to attenuate PRS performance in the present prospective setting. Consistent with this interpretation, predictive performance improved markedly in participants with epilepsy onset at ≤18 years of age, a subgroup expected to contain a greater proportion of genetically determined epilepsies.

Importantly, these findings should not be interpreted as evidence against a major genetic contribution to epilepsy. GGE is among the most heritable common neurological disorders, and the predominance of MR signals observed for GGE in our study further supports the strong influence of inherited susceptibility in this subtype. Rather, our results suggest that circulating proteomic biomarkers capture dynamic biological processes that are shared across genetically determined and acquired epilepsies and therefore provide complementary information to inherited genetic risk in population-based cohorts. This distinction between inherited susceptibility and dynamic disease-associated biology is likely to become increasingly important as multi-omic approaches are integrated into epilepsy risk prediction and precision medicine. Finally, although MR and colocalization analyses strengthened causal inference for several proteins, experimental validation will be required to confirm the biological mechanisms underlying the identified associations. In addition, independent replication in external prospective cohorts, including more diverse populations and larger numbers of clinically well-characterized epilepsy subtypes, will be essential to establish the generalizability and clinical utility of the proposed molecular risk scores.

By extending prospective plasma proteomics with metabolomics and human genetic analyses, our study demonstrates that prospective multi-omic profiling captures complementary dimensions of epilepsy biology that are not fully reflected by inherited genetic susceptibility alone. Circulating proteomic signatures provide the strongest predictive information, metabolomic profiles reveal systemic metabolic alterations preceding disease onset, and human genetics identifies a subset of proteins with evidence supporting causal involvement in epilepsy. Together, these findings establish a framework that combines prediction with mechanistic prioritization, advancing molecular risk stratification and potential therapeutic target discovery for epilepsy while illustrating the broader potential of prospective multi-omic profiling for precision medicine in complex neurological disorders.

## Data Availability

Access to the UKB data, including the proteomics and genetic datasets, is available to approved researchers upon application through the UKB access procedures.

## Acknowledgements

This study utilized data from the UKB through application number 73507. Some of the computational work was performed using the High-Performance Computing cluster at the University of Luxembourg (https://hpc.uni.lu/).

## Contributors

EH and PM conceptualised the study. EH developed the methodology, performed the formal analysis and data curation, prepared the visualisations, and wrote the original draft of the manuscript. PM supervised the study and acquired funding. EH and PM accessed and verified the underlying data. MEO, PP, HL, CL, DL, and PM critically reviewed and edited the manuscript. All authors read and approved the final version of the manuscript.

