## Supplementary Figures for "Prospective multi-omic profiling identifies predictive and genetically supported molecular determinants of incident epilepsy"

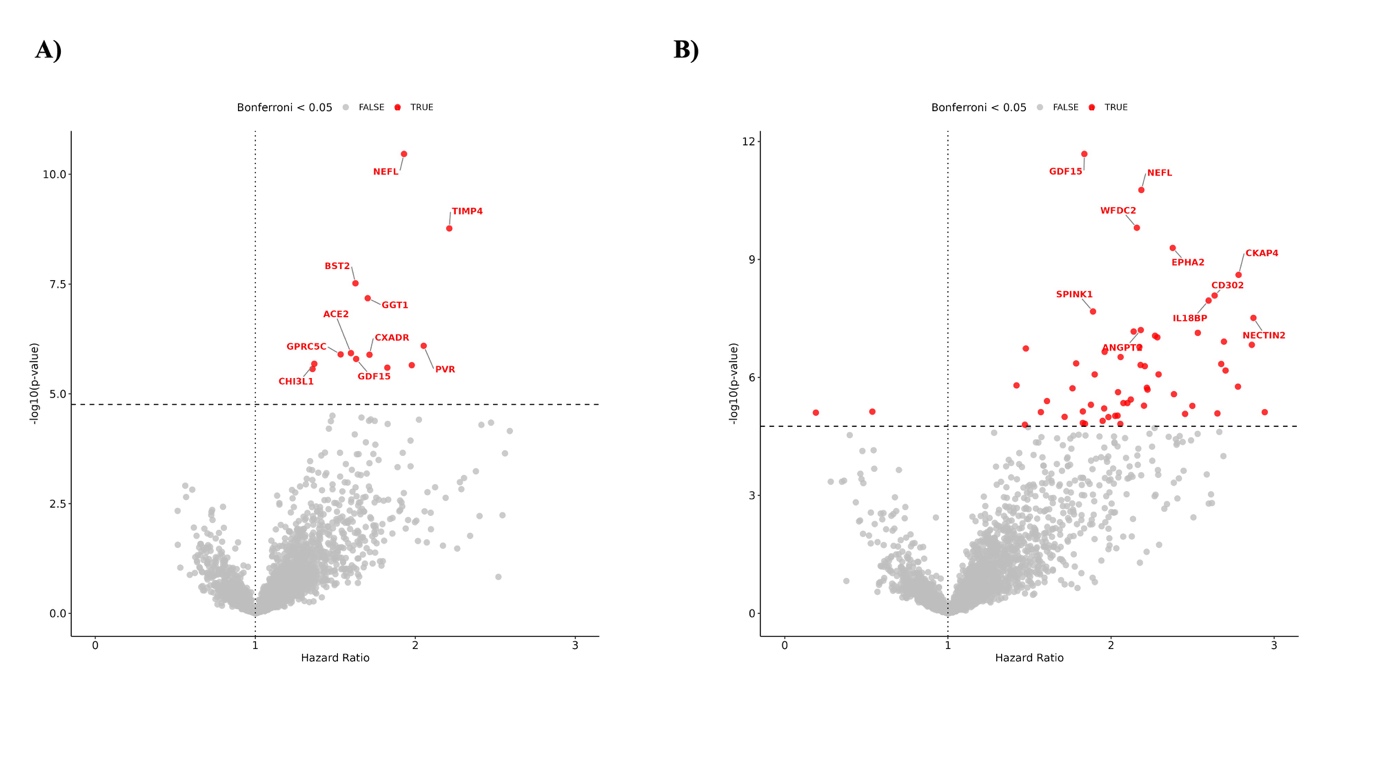

**Supplementary Figure S1: Plasma proteomic associations with incident epilepsy stratified by sex.** Volcano plot summarizes the results of the Cox proportional hazards regression models for plasma proteomic associations with incident epilepsy: A) males, B) females.

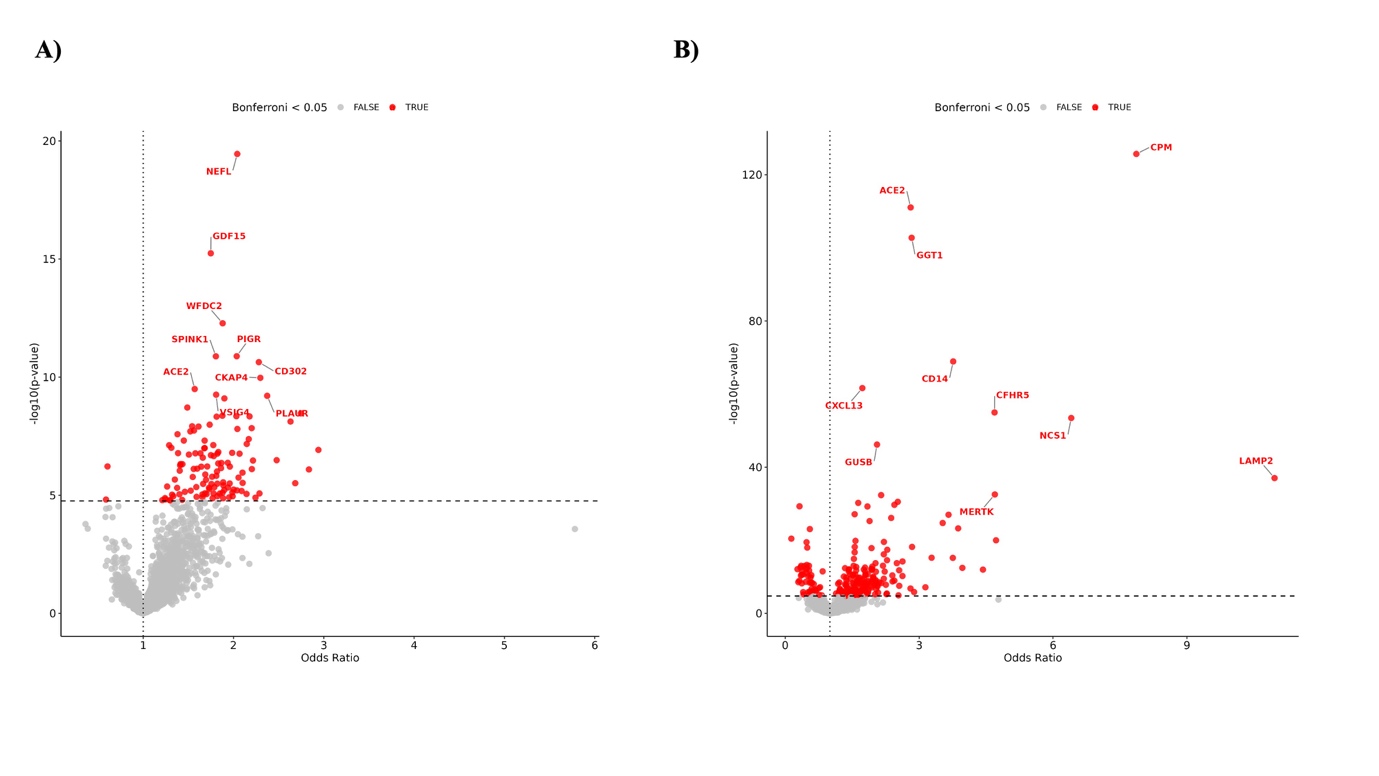

**Supplementary Figure S2: Plasma proteomic associations with incident and prevalent epilepsy.** Volcano plot summarizes the results of plasma proteomic associations: A) incident cases, B) prevalent cases.

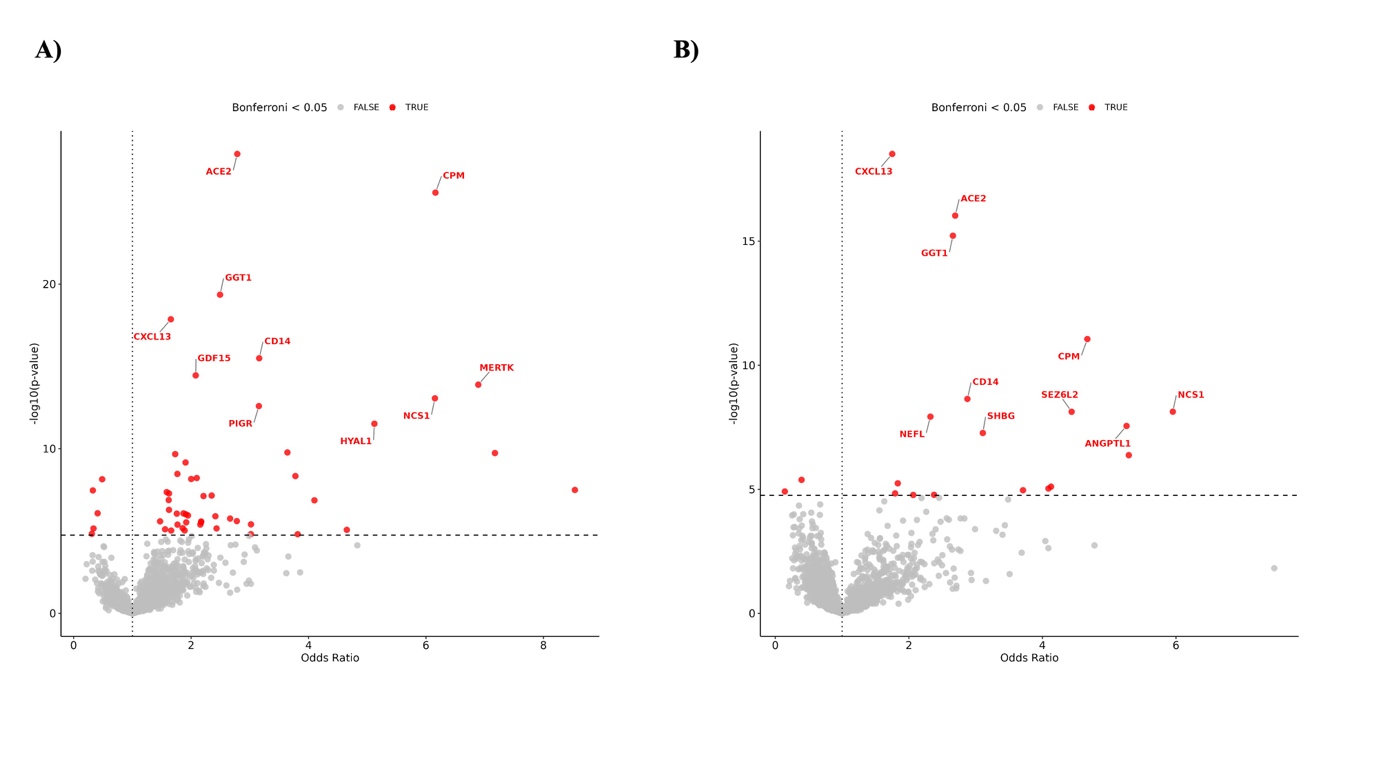

**Supplementary Figure S3: Plasma proteomic associations with genetic generalized epilepsy (GGE) and focal epilepsy (FE).** Volcano plot summarizes the results of the logistic regression models for plasma proteomic associations with A) GGE, B) FE.

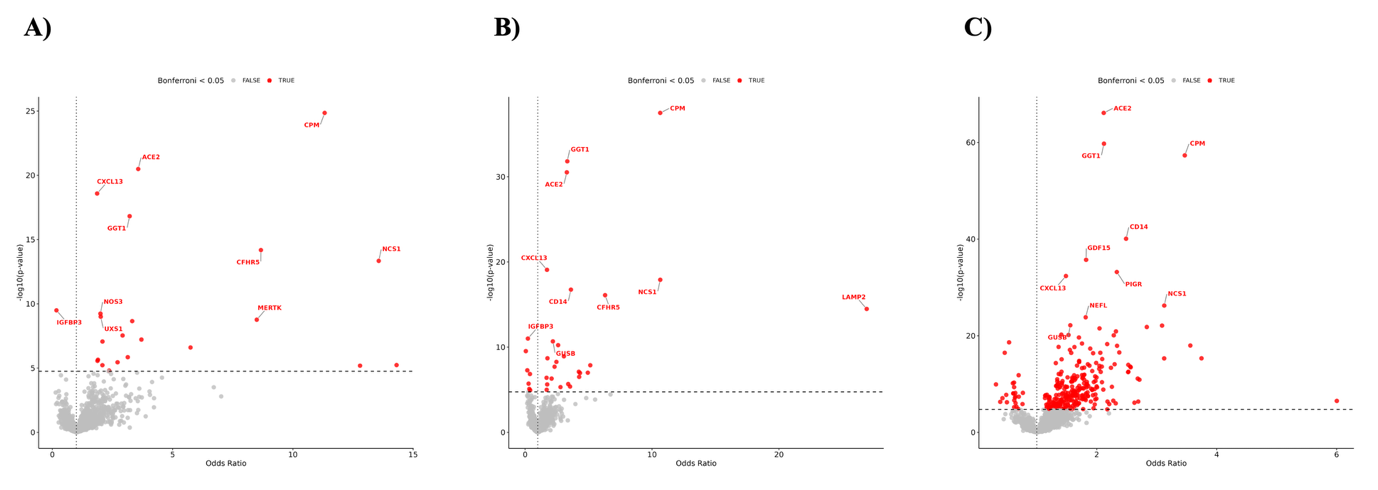

**Supplementary Figure S4: Plasma proteomic associations in epilepsy cases diagnosed <10 years and 10-18 years.** Volcano plot summarizes the results of the logistic regression models for plasma proteomic associations with cases diagnosed A) <10 years, B) 10-18 years, C) >18 years.

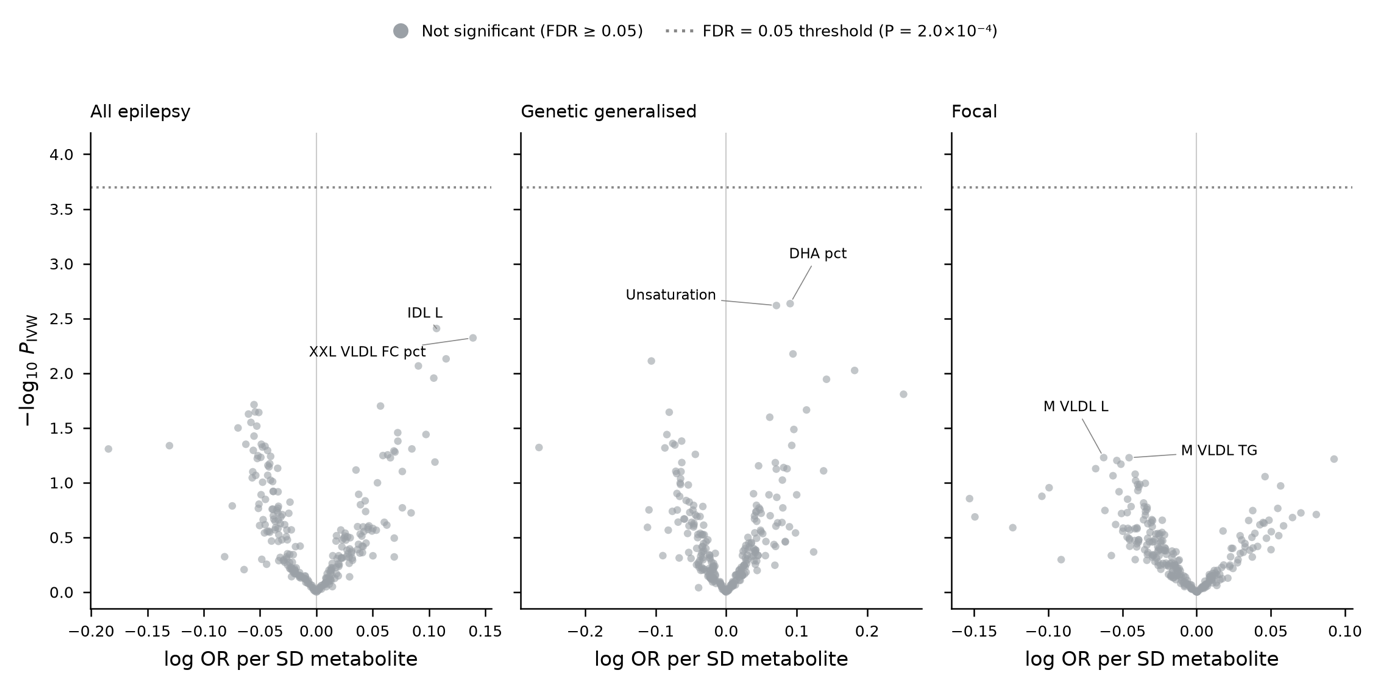

**Supplementary Figure S5: Metabolite-epilepsy Mendelian randomization.** Volcano plot summarizes the results of proteome-wide Mendelian randomization of metabolite levels against all epilepsy, genetic generalized epilepsy, and focal epilepsy.

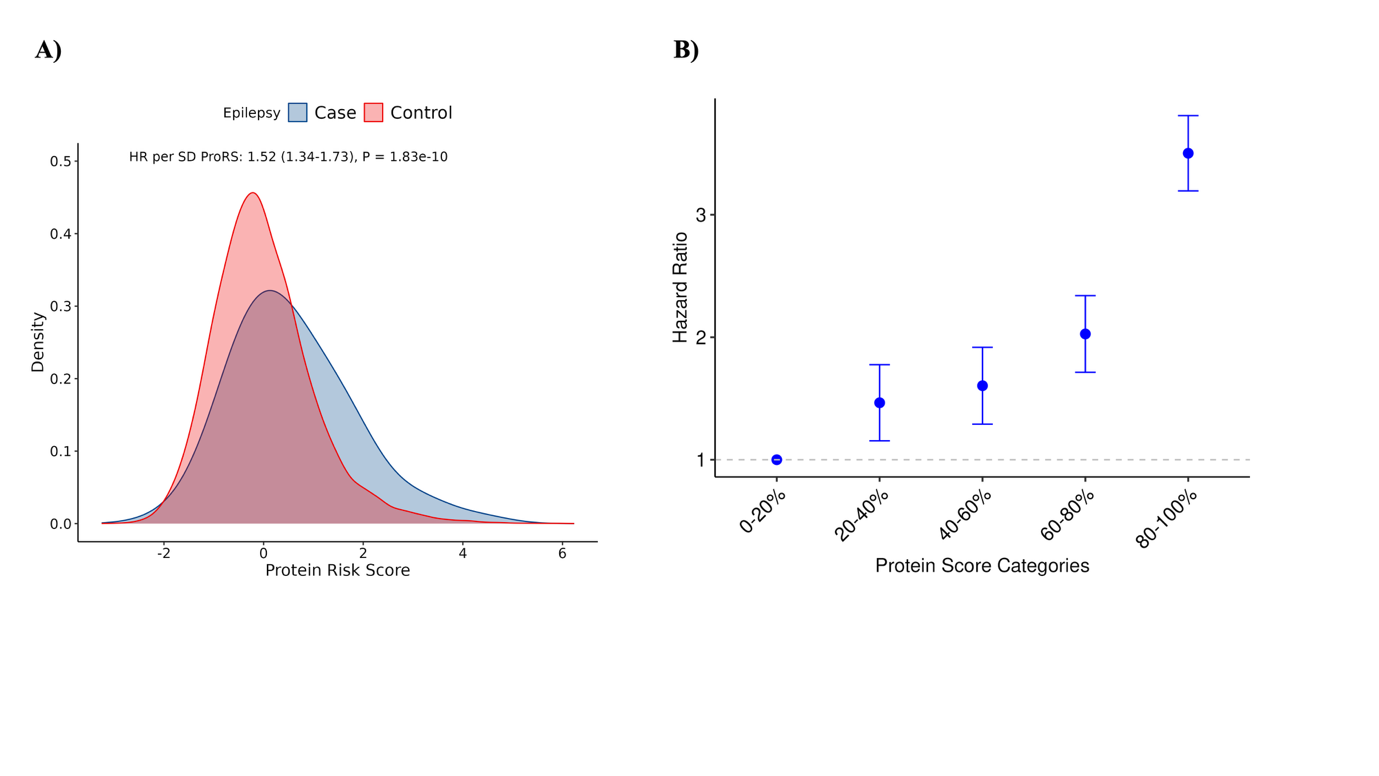

**Supplementary Figure S6: Protein risk score distribution and hazard ratio by score groups.** A) Density plot displays distribution of the ProRS for epilepsy cases (blue) and controls (red), illustrating the separation of scores between the two groups. B) Hazard ratio for incident epilepsy across different groups of the ProRS. The 0-20% group serving as the reference group (Hazard Ratio = 1). Error bars indicating the 95% confidence intervals.
